# Early Humoral Response Correlates with Disease Severity and Outcomes in COVID-19 Patients

**DOI:** 10.1101/2020.09.21.20198309

**Authors:** Anwar M Hashem, Abdullah Algaissi, Sarah A Almahboub, Mohamed A Alfaleh, Turki S Abujamel, Sawsan S Alamri, Khalid A Alluhaybi, Haya I Hobani, Rahaf H AlHarbi, Reem M Alsulaiman, M-Zaki ElAssouli, Sharif Hala, Naif K Alharbi, Rowa Y Alhabbab, Ahdab A AlSaieedi, Wesam H Abdulaal, Abdullah Bukhari, Afrah A AL-Somali, Fadwa S Alofi, Asim A Khogeer, Arnab Pain, Almohanad A Alkayyal, Naif AM Almontashiri, Ahmad Bakur Mahmoud, Xuguang Li

## Abstract

The Coronavirus Disease 2019 (COVID-19), caused by the novel SARS-CoV-2, continues to spread globally with significantly high morbidity and mortality rates. Immunological surrogate markers, in particular antigen-specific responses, are of unquestionable value for clinical management of patients with COVID-19. Here, we investigated the kinetics of IgM, IgG against the spike (S) and nucleoproteins (N) proteins and their neutralizing capabilities in hospitalized patients with RT-PCR confirmed COVID-19 infection. Our data show that SARS-CoV-2 specific IgG, IgM and neutralizing antibodies (nAbs) were readily detectable in almost all COVID-19 patients with various clinical presentations. Notably, anti-S and -N IgG, peaked 20-40 day after disease onset, and were still detectable for at least up to 70 days, with nAbs observed during the same time period. Moreover, nAbs titers were strongly correlated with IgG antibodies. Significantly higher levels of nAbs as well as anti-S1 and N IgG and IgM antibodies were found in patients with more severe clinical presentations, patients requiring admission to intensive care units (ICU) or those with fatal outcomes. Interestingly, lower levels of antibodies, particularly anti-N IgG and IgM in the first 15 days after symptoms onset, were found in survivors and those with mild clinical presentations. Collectively, these findings provide new insights into the characteristics and kinetics of antibody responses in COVID-19 patients with different disease severity.

## Introduction

The Coronavirus Disease 2019 (COVID-19) has first emerged in Wuhan, China as atypical pneumonia in a cluster of patients before it spread globally, causing a major pandemic that has affected more than 30 million people with around 950,000 deaths as of September 15, 2020 (WHO). The causative agent of COVID-19 was identified to be a novel betacoronavirus (beta-CoV) now known as severe acute respiratory syndrome coronavirus 2 (SARS-CoV-2) [1, 2]. Similar to other human CoVs, such as the Middle East respiratory syndrome coronavirus MERS-CoV and SARS-CoV, a zoonotic origin of SARS-CoV-2 was suggested but yet to be confirmed [2–4]. SARS-CoV-2 can infect individuals from different age groups and causes a wide spectrum of disease manifestations ranging from asymptomatic, mild, moderate to severe symptoms with possible fatal outcomes [5–7]. While the reasons of this wide range disease presentations could be attributed to many factors such as age, sex, pre-existing comorbidities, host genetics as well as other factors, host immune response is a key aspect in determining infection outcomes [7–10]. Specifically, specific humoral immune responses to the virus could dictate the disease outcomes; however, the early dynamics of antibody responses, including neutralizing antibodies (nAbs), in COVID-19 patients with different clinical presentations is still not well-characterized. Such information can help our understanding of the nature of COVID-19 infection, guide patient management as well as aid in the development and evaluation of therapeutics and vaccines. Indeed, the dynamics of early viral-specific antibody responses could influence the progression of several other viral infections including HIV, Influenza and Ebola [11–13].

One of the SARS-CoV-2 proteins capable of inducing robust immune response is the viral spike (S)protein. S protein is comprised of S1 and S2 subunits, with the former known to mediate binding to Angiotensin-converting enzyme 2 (ACE2) receptor on host cells and the latter being involved in viral-host membranes fusion [14, 15]. As ACE2 is the main receptor, nAbs mainly target the S1 subunit. As such, quantitative determination of antibodies against the S protein is widely used to characterize the antibody responses in COVID-19 patients. A recent study analyzed antibody responses in a small cohort of COVID19 patients found a strong association between the magnitude of anti-S antibody response and patient survival [16].

Another important viral protein capable of inducing strong immune response is the abundantly expressed nucleocapsid (N) protein [17, 18]. However, antibodies elicited towards the N protein are not neutralizing and might not provide protection against infection as shown in SARS-CoV infection model [19]. Interestingly, there is a report linking higher anti-N antibody response to severe and fatal outcomes [20]. These data suggest that early humoral immune responses should be further investigated.

In the past few months, several studies have focused on studying the humoral response against SARS-CoV-2 in COVID-19 patients, with antibodies against S1 and N antigens found to emerge as early as one week following disease onset and persist for at least three month after infection [18, 20–24]. Here, we studied the kinetics of SARS-CoV-2 specific antibodies to S1 and N viral proteins in blood samples collected between 4 to 70 days post-symptoms onset from a cohort of 87 COVID-19 patients with different disease presentations (i.e. mild, moderate or severe) or outcomes (i.e. survival vs death). As a control, we included serum samples collected before the emergence of SARS-CoV-2. Our data show that most of the COVID-19 patients with different disease categories were able to elicit specific anti-S1 and N IgG and IgM antibodies as well as nAbs that were well-maintained through the period of observation (70 days). We found that the levels of nAbs were strongly correlated with anti S1-IgG antibody response. Additionally, our data show a trend of high levels of anti-N IgG and IgM antibodies in COVID-19 patients based on disease severity, particularly in the first two weeks after the onset of symptoms.

## Material and methods

### Human subjects

Signed informed consent forms were obtained from all patients as per institutional ethical approvals obtained from the Unit of Biomedical Ethics in King Abdulaziz University Hospital (Reference No 245-20), the Institutional Review Board at the Ministry of Health, Saudi Arabia (IRB Numbers: H-02-K-076-0320-279 and H-02-K-076-0420-285), and the Global Center for Mass Gatherings Medicine, Saudi Arabia (GCMGM) (No. 20/03A).

### Samples

Between March 31^st^ to June 28^th^, 2020, 240 blood samples from 87 patients with confirmed SARS-CoV-2 infection by real-time RT-PCR were collected between days 4 and 70 post-symptoms onset. Patients were enrolled from two hospitals in Saudi Arabia; Ohoud hospital in Madinah (n=43 patients), and King Abdullah Medical Complex (KAMC) in Jeddah (n=44 patients). Longitudinal blood samples were collected from mild cases (n=46 patients, 128 samples), moderate cases (n=13 patients, 39 samples) and severe cases (n=28 patients, 73 samples) for serological analysis. The disease severity was categorized based on Saudi MOH guidelines. Patients with mild symptoms and no oxygen requirements or pneumonia on chest X-ray were classified as mild cases. Patients with respiratory symptoms and lung infiltrates in less than 50% of the lung field were considered as moderate cases. Patients who had one or more of the following symptoms (respiratory rate more than 30 breaths/minutes, blood oxygen saturation <93%, PaO2/Fio2 <300, lung infiltrates in more than 50% of the lung field within 24-48 hours) were considered as severe cases. Demographics, clinical data and disease outcomes were retrieved from the medical records. Sera (n=50) from healthy donors collected before COVID-19 pandemic were included as negative controls.

### Cells

Baby Hamster kidney BHK-21/WI-2 cell line (Kerafast, EH1011) and African Green monkey kidney-derived Vero E6 cell line (ATCC, 1586) were cultured in Dulbecco’s modified essential medium (DMEM) contained 100 U/ml of penicillin, and 100 μg/ml of streptomycin and supplemented with 5 and 10% fetal bovine serum (FBS) in a 5% CO_2_ environment at 37°C.

### Indirect ELISA

Recombinant SARS-CoV-2 S1 subunit (amino acids 1–685) was purchased commercially (Sino Biological, China). Recombinant SARS-CoV-2 N protein was expressed and purified in-house as previously described [18]. Indirect S1-based or N-based ELISA were performed for the detection of specific IgG and IgM as previously described [18]. Briefly, Recombinant S1 and N proteins were used to coat 96-well high binding ELISA plates (Greiner Bio One, Monroe, NC) at 1□μg/ml and 4□μg/ml in phosphate-buffered saline (PBS) with 50 μl per well, respectively. After overnight incubation at 4°C, plates were washed with PBS containing 0.05% tween-20 (PBS□T) and blocked with 5% skim milk in PBS-T buffer at 37°C for 1□h. Plates were then washed and incubated with serum samples diluted at 1:100 in PBS□T□with□5% milk for 1□h at 37°C. After another washing, plates were incubated with HRP□conjugated goat anti□human IgG (H□+□L) or IgM antibodies (Jackson ImmunoResearch, West Grove, PA) for 1 h, washed again, and incubated with TMB (3,3’,5,5’-tetramethylbenzidine) substrate (KPL, Gaithersburg, MD) at room temperature for 30□min. The reaction was stopped by 100 μl per well of 0.16 M sulfuric acid, and absorbance was measured at 450 □nm using the ELx808™ Absorbance Microplate Reader (BioTek, Winooski, VT). Cutoff values for these ELISA were previously determined as 0.17 for IgG S1-ELISA, 0.3 for IgM S1-ELISA, 0.4 for IgG N-ELISA, and 0.55 for IgM N-ELISA [18].

### Generation of rVSV-ΔG/SARS-2-S*-luciferase pseudovirus

The pseudovirus was produced and titrated as previously described [25]. Briefly, a T-175 tissue culture flask of BHK21/WI-2 cells were transfected with 46 μg of pcDNA expressing codon-optimized full-length SARS-CoV-2 S protein (GenBank accession number: MN908947) using Lipofectamine ™ 2000 transfection reagent (Invitrogen). After 24 h post-transfection, cells were infected with rVSV-ΔG/G*-luciferase (Kerafast, EH1020-PM) at a multiplicity of infection (m.o.i.) of 4. The virus inoculation was removed after 1 h and the cells were washed twice with 12 ml PBS. To completely remove any excess amount of the rVSV-ΔG/G*-luciferase, 15 ml of DMEM containing rabbit polyclonal anti VSV-G antibody were added to the cells monolayer and incubated for 24 h at 37°C in 5% CO_2_ humidified incubator. Supernatant containing the generated pseudovirus was harvested 24 h post-infection and the virus titer was determined by measuring luciferase activity from 2-fold serially diluted pseudovirus on Vero E6 cells as previously described [25]. The titer of pseudovirus was expressed as a relative luciferase unit (RLU).

### Pseudovirus neutralization assay

Neutralization assay was performed as previously reported [25]. In brief, Vero E6 cells were seeded in 96-well plates (white plate with clear bottom, COSTAR) at 2 × 10^4^ cells/well and incubated overnight at 37°C in 5% CO_2_ humidified incubator. Human sera were heat-inactivated at 56°C for 30 min, and diluted as half log serial dilutions with an initial dilution of 1:20 in DMEM containing 5% FBS. Each serum dilution was mixed with diluted pseudovirus that yields 2 × 10^4^ RLU and incubated at 37°C, 5 % CO_2_ for 1 h in duplicates. Then, a 100 μl of the pseudovirus–serum mixtures were transferred onto Vero E6 cell monolayers and incubated at 37°C in a 5% CO_2_ humidified incubator for 24 h. Cell only control (CC) and virus control (VC) were included with each assay run. After 24 h, cells were lysed, and luciferase activity was measured using the Luciferase Assay System (Promega) according to the manufacturer’s instructions. The inhibition of luciferase activity by each dilution of the serum sample was determined as follows: 100 – [(mean RLU from each sample – mean RLU from CC) / (mean RLU from VC – mean RLU from CC) x 100]. Median Inhibitory Concentration (IC_50_) neutralization titers were determined using four-parameter logistic (4PL) curve in GraphPad Prism V8 software (GraphPad Co.).

### Statistical analysis

Statistical analyses and graphical presentations were conducted with GraphPad Prism version 8.0 software (Graph-Pad Software, Inc., CA, USA) and R (version 4.0.0). Statistical analysis of the different severity groups (mild, moderate and severe) were conducted using one-way analysis of variance with Tukey post-hoc test to adjust for multiple comparisons between groups. Differences of antibody responses between disease outcomes (death vs survival) and between ICU and non-ICU patient groups were determined by Student’s t test. Pearson’s correlation coefficient was used to assess the relationship between anti-S1 and anti-N IgG and IgM from ELISA and nAbs titers as well as between anti-S1 IgG and anti-N IgG, and anti-S1 IgM and anti-N IgM. The scatterplot smoothing lines of IgG, IgM or nAbs titers overtime were generated using locally estimated scatterplot smoothing (LOESS) regressions with ribbons depicting the 95% confidence intervals using the ggplot package in R. All values are depicted as mean ± SD and statistical significance is reported as *, P≤0.05; **, P≤0.01; ***; P≤0.001; and ****, P≤0.0001.

## Results

### SARS-CoV-2 induces sustained antibody response in patients despite disease severity

To study the kinetic of SARS-CoV-2 specific antibody responses over time in COVID-19 patients with different disease severity, we collected a total of 240 serial blood samples from 87 patients with real time RT-PCR confirmation of SARS-CoV-2 infection between days 4 and 70 post-symptom onset. The COVID-19 cohort included mild (n=46 patients, 128 samples), moderate (n=13 patients, 39 samples) and severe cases (n=28 patients, 73 samples).

We initially analyzed the overall nAbs and anti-S1 and N IgG and IgM antibody responses in our COVID-19 cohort. Almost all COVID-19 patients in our study produced and maintained high titers of nAbs (**Figure 1a**) and mounted a high level of IgG and IgM antibodies to SARS-CoV-2 S1 (**Figure 1b**) and N proteins (**Figure 1c**) despite their disease severity (**Supplementary Figure 1**). Furthermore, the observed high levels of nAbs and anti-S1 and N IgG peaked between days 20 and 40 and were maintained up to 70 days post-disease onset, whereas IgM levels waned overtime. Nonetheless, there were some variations in the magnitude of response between patients within each category. While almost all cases especially moderate ones seroconverted during the course of samples collection and eventually produced robust nAbs, IgG and IgM antibodies, few exceptions were observed in mild and severe categories. Specifically, no detectable anti-S1 and weak anti-N IgG antibodies were found in one mild case in addition to loss of nAbs, coinciding with reduced level of anti-S1 IgG by day 60, was observed in another mild case although high levels of anti-N IgG were detected (**Figures 1d-f**). Furthermore, in two severe cases no nAbs or anti-S1 IgG at days 20 and 30 were found, although low levels of anti-N IgG and IgM and anti-S1 IgM were induced (**Figures 1d-f**). Taken together, these data show that SARS-CoV2 S1 and N specific antibodies were readily detectable in high levels that were sustained for up to 70 days in most COVID-19 cases, with nAbs being induced in all clinical categories of COVID-19 patients.

**Figure 1.**
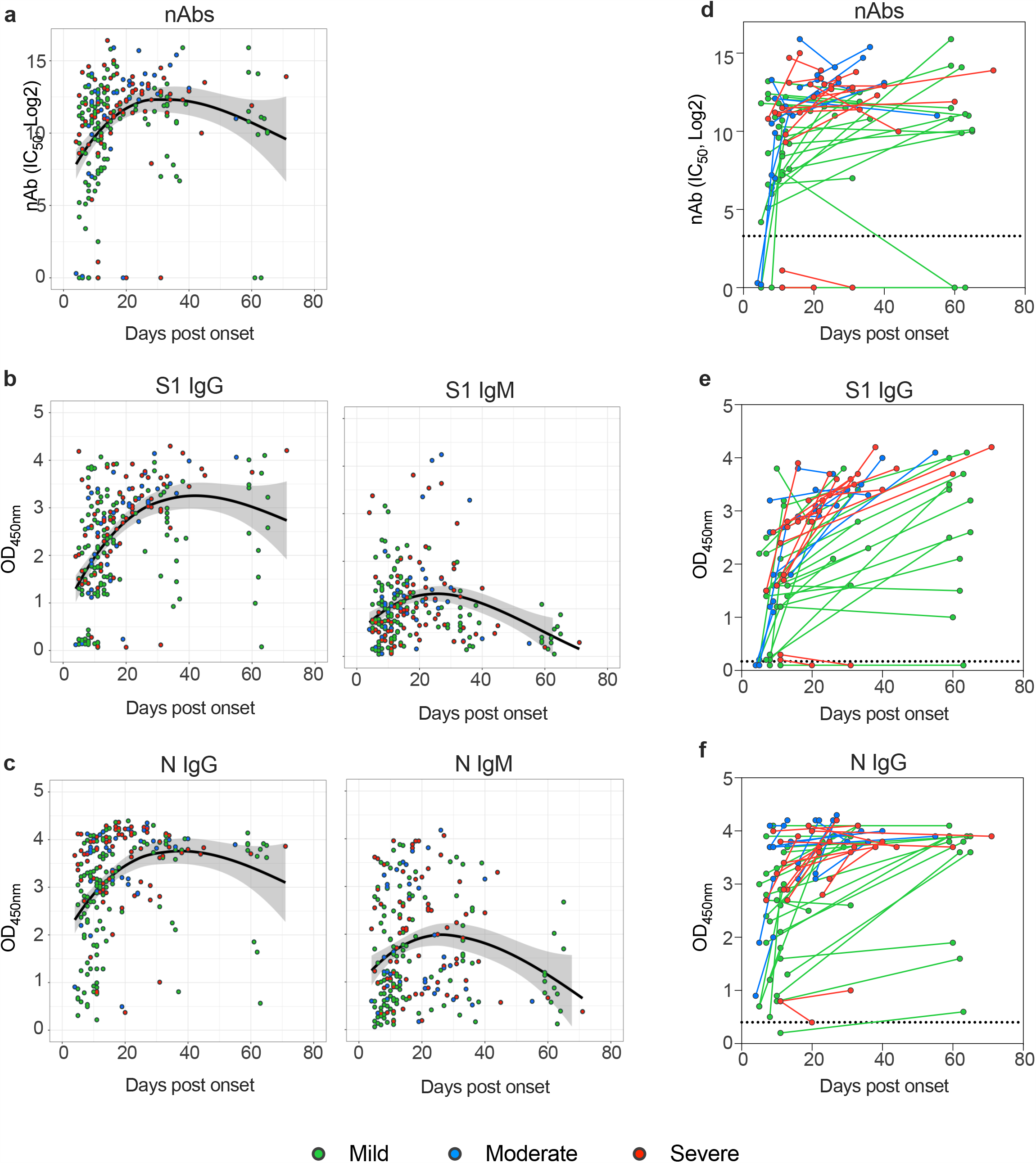
Kinetics of SARS-CoV-2 specific antibodies over time based on disease severity. (**a**) nAbs titers (IC_50_) and OD values of (**b**) S1-specific IgG and IgM and (**c**) N-specific IgG and IgM are plotted against the days post disease onset. The line and the ribbon show the mean expected from a LOESS regression model with 95% confidence interval. The colored circles indicate disease severity. OD = optical density. Changes in (**d**) nAbs titers (IC_50_) and OD values of (**e**) S1-specific IgG and (**f**) N-specific IgG for each patient are shown over time. Data from first sample and last collected sample are shown for patients with samples that at least 7 days apart.

### Both S1 and N-specific IgG antibodies correlate with each other and with nAbs in COVID-19 patients

Next, we examined the correlation between S1-IgG, N-IgG, S1-IgM and N-IgM antibodies and nAbs titers based on disease severity. As shown in **Figure 2a**, in all disease categories, levels of anti-S1 and anti-N IgG independently had a significant and strong correlation with nAbs levels (r=0.9005, p<0.0001 and r=0.8899, p<0.0001, respectively), while moderate but significant correlation was also observed between nAbs and anti-S1 IgM (r=0.5785, p<0.0001) and anti-N IgM (r=0.5633, p<0.0001).

**Figure 2.**
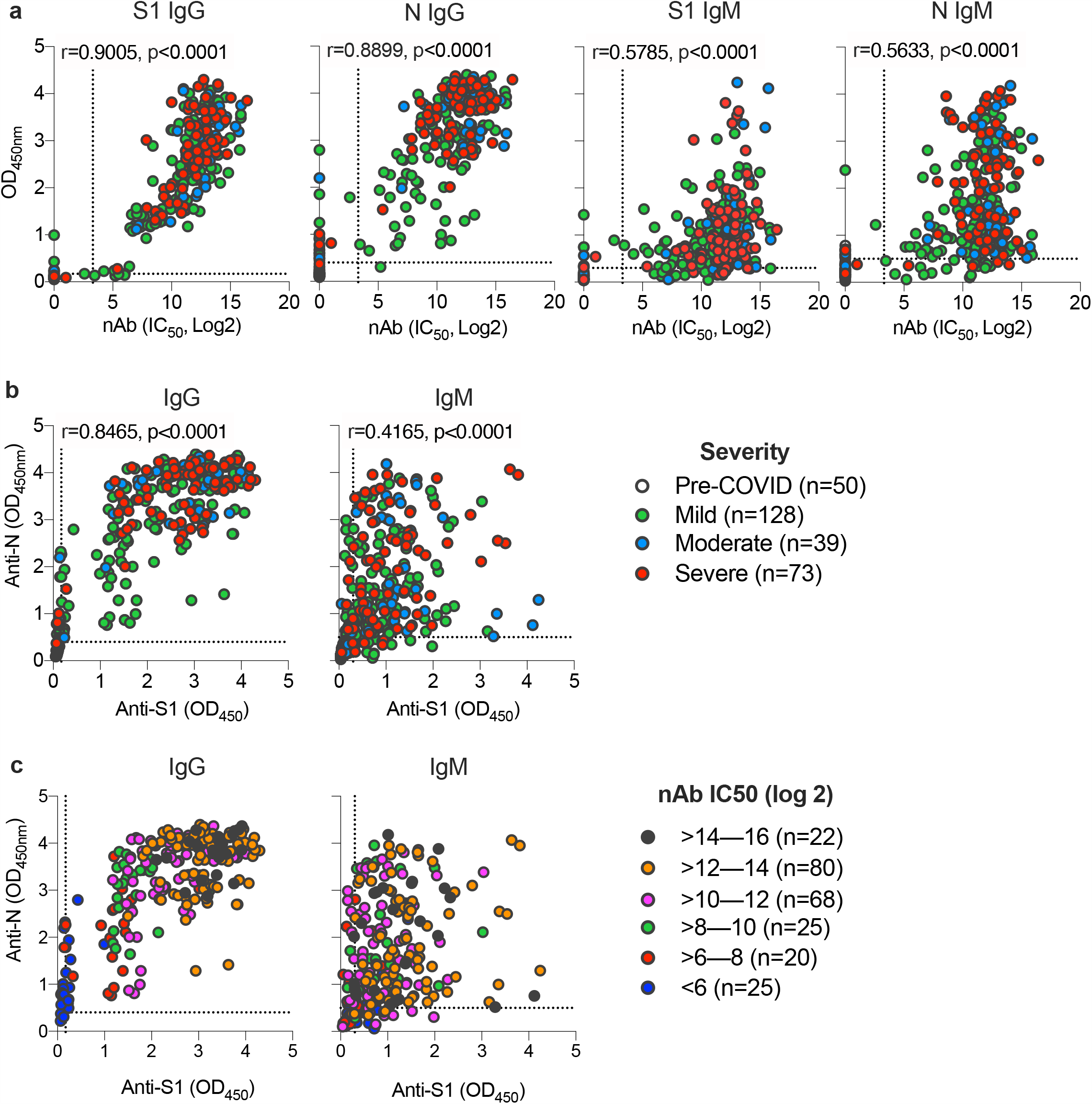
Relationship between S1 and N-specific IgG and IgM and neutralizing titer based on disease severity. (**a**) Pearson’s correlation between S1-IgG, N-IgG, S1-IgM and N-IgM and neutralizing titer based on disease severity. Scatter plots were generated using OD values (y-axis) versus IC_50_ (x-axis) from each individual serum sample. Colored circles indicate disease severity.; (**b**) Pearson’s correlation between IgG response against S1 and N proteins and IgM response against S1 and N proteins based on disease severity. Scatter plots were generated using anti-N OD values (y-axis) versus anti-S1 OD values (x-axis) from each individual serum sample. Colored circles indicate disease severity.; (**c**) Pearson’s correlation between IgG response against S1 and N proteins and IgM response against S1 and N proteins based on neutralizing titer. Colored circles indicate neutralizing titers (IC_50_). The dotted lines represent the cut-off of each assay. OD = optical density.

Interestingly, anti-N IgG antibodies seem to be higher in COVID-19 patients with severe infection than moderate or mild infections, while anti-S1-IgG antibodies appeared to be similar across all disease categories (**Figures 2a and 2b**). Furthermore, while both anti-S1 and anti-N IgG significantly and strongly correlate with each other (r=0.8465, p<0.0001), IgM against S1 and N proteins seem to have lower correlation (r=0.4165, p<0.0001) (**Figure 2b**). We also found that high levels of nAb titers correlates with higher levels of IgG response against both S1 and N proteins (**Figure 2c**). These data indicate that anti-S1 and anti-N IgG levels strongly correlate with each other and with nAbs titers, and higher levels of anti-N responses could be associated with more severe disease.

### Disease severity impacts antibody responses against SARS-CoV-2

To study the correlation between the disease severity and specific antibodies response in more details, we compared antibody levels in the different COVID-19 disease categories. As shown in **Figure 3a**, COVID-19 patients who developed severe or moderate infections mounted significantly higher overall nAbs levels compared to mildly infected individuals. Additionally, S1-IgG antibodies were significantly higher in severe cases than the mild ones. Importantly, levels of anti-N IgG and IgM were significantly associated with disease severity and higher levels were induced in moderate and severe cases compared to mild infections. When stratifying patients based on their need for ICU admission or based on infection outcome, we found that nAbs as well as anti-S1 and N antibodies (IgG and IgM) were significantly higher in those who required ICU or those with fatal outcomes (**Figures 3b and 3c**). These data clearly show a significant correlation between COVID-19 disease severity and the overall humoral immune responses.

**Figure 3.**
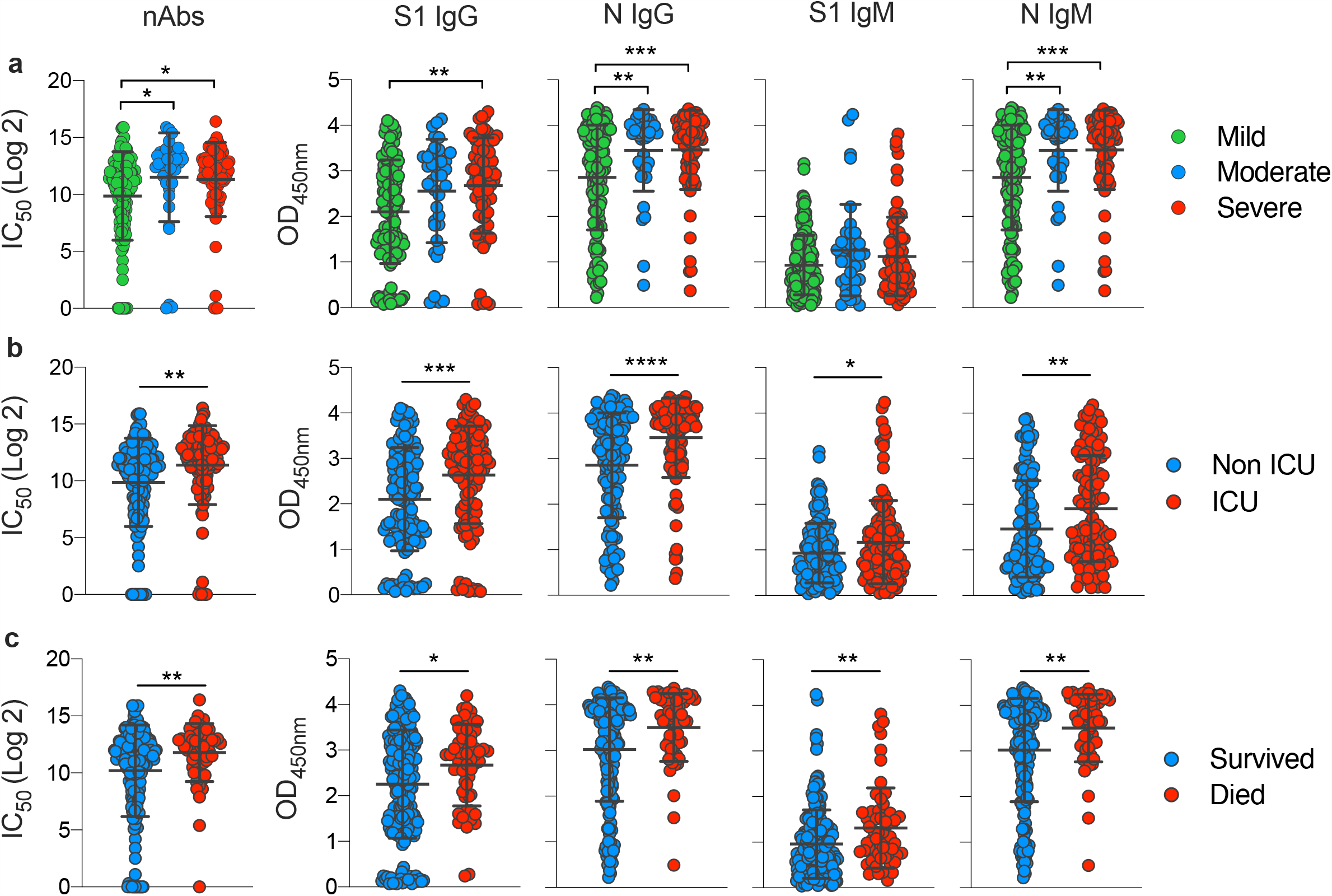
Overall impact of disease severity on antibody responses against SARS-CoV-2. Comparison of neutralizing titer (IC_50_) and OD values of S1-IgG, N-IgG, S1-IgM and N-IgM for (**a**) individuals from the different disease severity categories; (**b**) individuals who survived or died; and (**c**) individuals who required ICU admission or not. Statistical significance was determined using one-way analysis of variance with Tukey post-hoc test in (**a**) and using Student’s t test in (**b**) and (**c**). Mean with SD are shown, and statistical significance is represented as *, P≤0.05; **, P≤0.01; ***; P≤0.001; and ****, P≤0.0001.

To better understand whether disease severity could impact the kinetic of antibody response in patients, we analyzed antibody responses in our COVID-19 cohort over time during the observation period. While almost all patients produced nAbs and anti-S1 antibodies with no significant difference over time except for anti-S1 IgG at late time points (**Figure 4a**), individuals with severe symptoms appeared to mount a significantly higher anti-N IgG and IgM during the first 15 days (acute phase) post-disease onset compared with individuals with milder disease (**Figure 4a**). Similarly, higher levels of anti-N IgG and IgM during the acute phase were more significantly associated with the need for ICU admission (**Figure 4b**). Furthermore, anti-S1 antibodies seem to be sustained at a significantly higher levels after day 30 post-symptoms onset in ICU patients compared to non-ICU patients. Comparing the kinetics of antibody response in COVID-19 patients who had fatal outcomes to those who survived the infection also showed that early induction of anti-N IgG and IgM during the first 15 days post-disease onset is indicative of fatal outcomes (**Figure 4c**). Additionally, we observed a significantly higher nAbs and anti-S1 IgG induction during the first 15 days in those who succumbed to infection compared to those who survived (**Figure 4c**). Taken together, these observations revealed that type and dynamics of specific antibodies responses could be associated with disease outcome.

**Figure 4.**
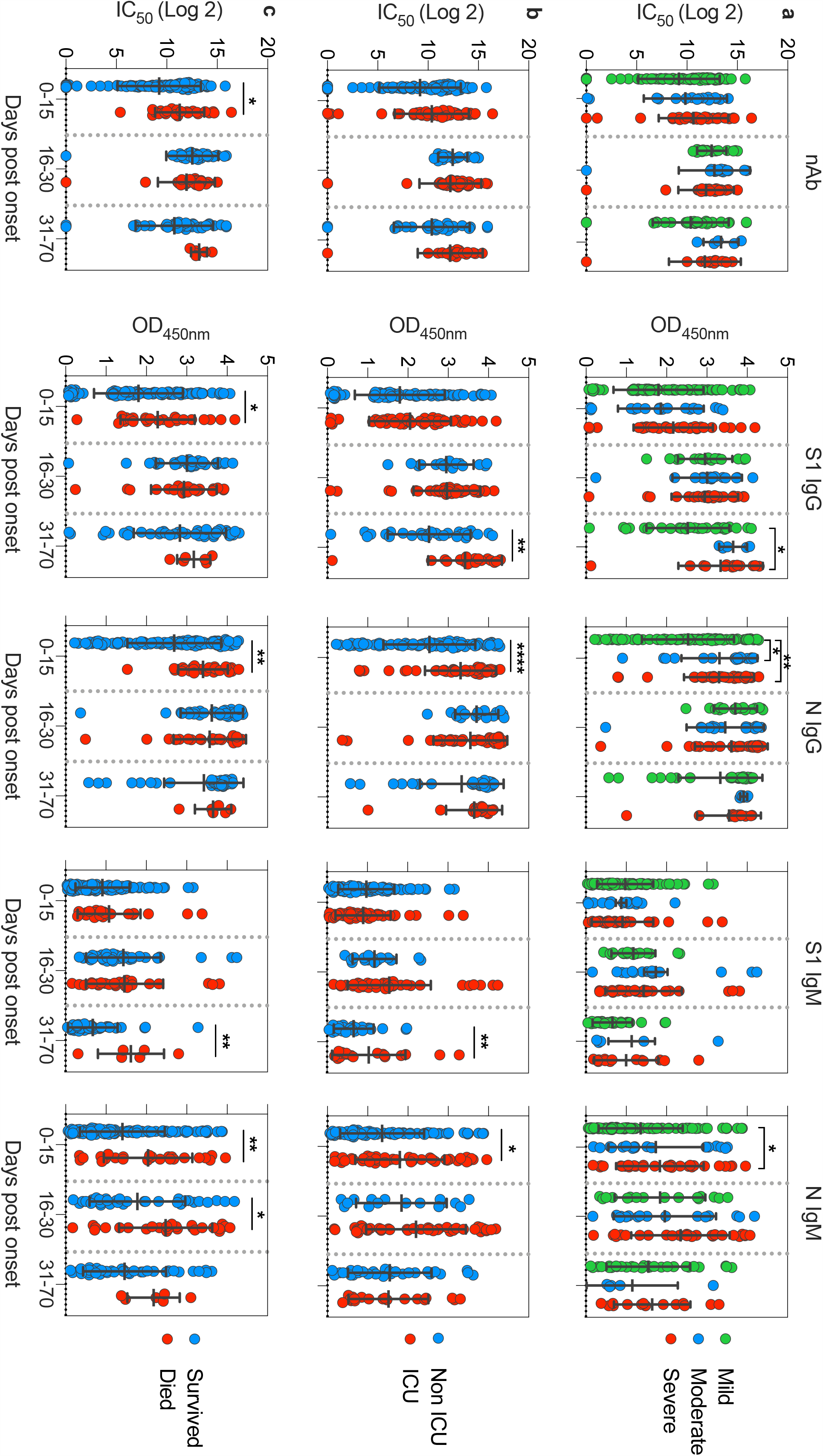
Impact of disease severity on kinetics of antibody responses against SARS-CoV-2. Comparison of neutralizing titer (IC_50_) and OD values of S1-IgG, N-IgG, S1-IgM and N-IgM for (**a**) individuals from the different disease severity categories; (**b**) individuals who survived or died; and (**c**) individuals who required ICU admission or not. Data are shown based on time post disease onset. Statistical significance was determined using one-way analysis of variance with Tukey post-hoc test in (**a**) and using Student’s t test in (**b**) and (**c**). Mean with SD are shown, and statistical significance is represented as *, P≤0.05; **, P≤0.01; and ****, P≤0.0001.

## Discussion

As the COVID-19 global pandemic continues to spread, there is an urgent need to conduct in-depth analyses of immune responses in COVID-19 patients. Specifically, characterization of the type and kinetics of specific antibody responses would be crucial in advancing our understanding of the mechanisms underlying disease severity or outcome [5].

Thus far, there have been a number of studies on SARS-CoV-2 specific antibody responses in COVID-19 patients, but the interpretation of antibody responses data and association with disease severity remains not well understood. In this study we studied the characteristics and kinetics of SARS-CoV-2 specific antibody response (nAbs, IgG and IgM) in a series of serum samples collected from a total of 87 confirmed COVID-19 hospitalized patients over a period of 70 days post-symptoms onset. A total of 240 blood samples were collected from the cohort, which included mild cases (n=46 patients, 128 samples), moderate cases (n=13 patients, 39 samples) and severe cases (n=28 patients, 73 samples), in addition to pre-pandemic serum samples (50 samples). These serum samples were analyzed for specific IgG and IgM responses against SARS-CoV-2 S1 and N antigens, as well as for the nAbs levels using SARS-CoV-2 S pseudovirus-based assay.

Our data showed that anti-S1 and N IgG and IgM antibodies as well as nAbs were readily detectable across all COVID-19 disease categories and that these antibody responses can persist for at least up to 70 days post disease onset, which is the observation period in this study. We also observed that, on average, IgG antibodies for S1 and N were higher in magnitude than IgM antibodies with a peak time for both antibodies at around days 20 to 40 post infection. Moreover, IgG antibodies continued at a plateau level thereafter, while the IgM antibodies declined significantly after about 40 days post infection. Clearly, the finding that most COVID-19 patients in this study mounted high levels of sustained antibody responses is encouraging, consistent with some of the previous observations [16, 21, 24, 26, 27]. It is noteworthy that the observed decline in antibody response after about day 40 which was followed by a sustained high response until the end of the observation period should not raise concerns as infection usually leads to generation of SARS-CoV-2-specific memory B and T cells that can persist for long times and could provide protection against reinfection as recently demonstrated [28].

In our study we also show that the induction of high nAbs titers was strongly correlated with IgG response and in particular anti-S1 IgG. S1 subunit of the Spike protein mediates the binding with ACE2 receptor through the receptor binding domain (RBD). It has been shown that the RBD is the main target for most of the nAbs which may explain our finding [15, 29]. This finding may also suggest the utility of using serological testing on IgG antibodies as an alternative in the absence of neutralization assays. Moreover, nAbs as well as anti-S1 and N IgG and IgM antibodies were significantly higher in those who required ICU or those with fatal outcomes than non-ICU or those who survived the infection, suggesting that high antibody responses may be considered as a risk factor of severe and fatal outcomes.

Importantly, when we compared the kinetics of antibody responses across the COVID-19 disease severity spectrum, we found that anti-N IgG antibodies were more likely to correlate with disease severity than anti-S1 IgG, especially when mild and severe cases were compared. During the first 15 days, we noticed significantly higher levels of anti-N IgG and IgM in severe and moderate cases compared to mild infection and in ICU compared to non-ICU patients. Analysis of SARS-CoV-2 specific antibodies response in a cohort of COVID-19 patients in China showed a potential overall correlation between total antibodies (IgG and IgM) against the RBD and disease severity [30]. Interestingly, a recent study showed an opposite trend of increased IgG antibodies in milder cases compared to more severe COVID-19 patients [31]. However, this study focused on the correlation between antibodies levels and viral RNA shedding in patients and not necessarily disease severity. In our analysis, however, it was clear that early after disease onset, patients who had severe symptoms including those who were admitted to ICU and who had fatal outcomes showed significantly higher anti-N IgG and IgM antibodies. Considering that N protein is one of the highly expressed proteins, these high levels of anti-N antibodies could indicate a high virus replication in severely infected individuals compared to none severe patients. In addition, since anti-N antibodies are usually non-neutralizing, the high levels of these antibodies in severe patients may have contributed to the disease severity and immunopathology. It worth noting that presence of these kind of antibodies may raise concern of antibody-dependent disease enhancement which was reported in SARS patients and in feline coronavirus infections [32-34]. Nonetheless, these findings clearly indicate that early high anti-N antibodies could be an indicator of disease severity and could be used as a prognostic marker of disease outcomes, however, the implications and underlying mechanisms of such high levels of anti-N antibody responses require further studies.

In summary, our data indicates that COVID-19 patients can generate high levels of antibody responses that might last for a long period despite their disease severity. Our results also show that severe infections are associated with overall stronger anti-S1 and -N IgG and IgM antibodies as well as nAbs compared to none-severe cases. Importantly, significantly higher levels of antibodies, particularly anti-N IgG and IgM, were observed during the acute phase of the disease in severely ill cases which required ICU admission or succumbed to infection, suggesting they could be helpful prognostic markers of COVID-19. Nonetheless, additional studies are clearly needed to further understand the underlying molecular mechanisms by which antibody levels could impact disease course or prognosis.

## Data Availability

We confirm that all data related to the study are included in the manuscript

